# Effects of B.1.1.7 and B.1.351 on COVID-19 dynamics. A campus reopening study

**DOI:** 10.1101/2021.04.22.21255954

**Authors:** Kevin Linka, Mathias Peirlinck, Amelie Schäfer, Oguz Ziya Tikenogullari, Alain Goriely, Ellen Kuhl

## Abstract

The timing and sequence of safe campus reopening has remained the most controversial topic in higher education since the outbreak of the COVID-19 pandemic. By the end of March 2020, almost all colleges and universities in the United States had transitioned to an all online education and many institutions have not yet fully reopened to date. For a residential campus like Stanford University, the major challenge of reopening is to estimate the number of incoming infectious students at the first day of class. Here we learn the number of incoming infectious students using Bayesian inference and perform a series of retrospective and projective simulations to quantify the risk of campus reopening. We create a physics-based probabilistic model to infer the local reproduction dynamics for each state and adopt a network SEIR model to simulate the return of all undergraduates, broken down by their year of enrollment and state of origin. From these returning student populations, we predict the outbreak dynamics throughout the spring, summer, fall, and winter quarters using the inferred reproduction dynamics of Santa Clara County. We compare three different scenarios: the true outbreak dynamics under the wild-type SARS-CoV-2, and the hypothetical outbreak dynamics under the new COVID-19 variants B.1.1.7 and B.1.351 with 56% and 50% increased transmissibility. Our study reveals that even small changes in transmissibility can have an enormous impact on the overall case numbers. With no additional countermeasures, during the most affected quarter, the fall of 2020, there would have been 203 cases under baseline reproduction, compared to 4727 and 4256 cases for the B.1.1.7 and B.1.351 variants. Our results suggest that population mixing presents an increased risk for local outbreaks, especially with new and more infectious variants emerging across the globe. Tight outbreak control through mandatory quarantine and test-trace-isolate strategies will be critical in successfully managing these local outbreak dynamics.

## 1 Motivation

More than half a million cases of COVID-19 have been reported at United States colleges and universities since the beginning of the pandemic. Although many students, staff, and faculty have been immunized since vaccination became available in December 2020, more than 120,000 of all cases have occurred since January 2021 [19]. This is alarming, especially in view of the newly emerging variants of COVID-19 that are more infectious, and potentially more dangerous to the younger population [4]. American institutions of higher education remain faced with a difficult choice: maintaining campus life while exposing staff and students to a dangerous disease, or closing the campus at the risk of educational, mental, and financial disruption [17]. In most cases, university administrations soon realized that the risks of maintaining normal in-person learning during the pandemic were too grave and switched to an all-online instruction. Stanford University was the first major university to announce this transition on March 6, 2020. Within the following week, many colleges and universities sent home their students and by April 4, 2020, 1,400 university campuses had been closed [18]. While it became rapidly clear that campuses would not reopen for the remainder of the academic year, the next major question was to decide on possible reopening for the fall of 2020, especially in the light of the increase in prevalence among young adults. Indeed, young adults aged 20 through 29 years contributed substantially to the increase in COVID-19 infections across the southern United States during June 2020 [1].

One of the main risks associated with campus reopening is bringing students from high prevalence regions to campus and allowing them to mix with other students, possibly from low prevalence regions [8, 15]. Despite various mitigating strategies including testing, quarantine, contact tracing, facemask usage, and dedensification [16,28], the risk of creating an uncontrollable outbreak and spreading the disease to a large part of the campus population remains exceptionally high as demonstrated by the cautious reopening followed by the rapid closing of the University of North Carolina at Chapel Hill in August 2020 [27]. Despite some initial hopes of inviting student back to campus for the winter term, Stanford University remained closed to the majority of its students until the end of winter 2020.

The objectives of this study are twofold: First, we perform a retrospective study to evaluate the risks that would have been associated with the reopening of Stanford University in the spring, summer, and fall of 2020, and winter of 2021. Our analysis accounts for the full regional diversity of the student population by tracking their origin states and each state’s COVID-19 prevalence. We infer the critical parameters of the underlying network SEIR model using Bayesian analysis and estimate the number of students who would have been infected if the campus had fully reopened. Second, we complement our analysis by exploring the possible effect of variants on the overall disease dynamics.

## 2 Methods

### 2.1 Epidemiology modeling

We model the local epidemiology of the COVID-19 outbreak using an SEIR model [2, 5, 12, 21] with four compartments, the susceptible, exposed, infectious, and recovered populations, governed by the set of ordinary differential equations,

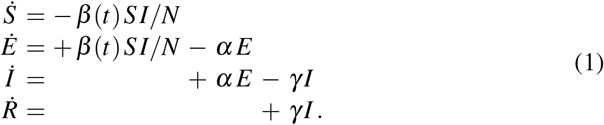

Here 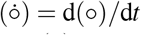denotes the time-derivative of the compartment (∘) and *N* = *S* + *E* + *I* + *R* is the total population. Three parameters govern the transition from one compartment to the next: the contact rate *β*, the latent rate *α*, and the infectious rate *γ*. They are the inverses of the contact period *B* = 1*/β*, the latent period *A* = 1*/α*, and the infectious period *C* = 1*/γ*. To keep the number of parameters manageable, we assume that latent rate *α* = 1*/*2.5 days^−1^ and the infectious rate *γ* = 1*/*6.5 days^−1^ are disease-specific for COVID-19, and constant in space and time [9, 11, 25]. To account for social behavioral changes during the course of the pandemic, we employ a dynamic contact rate *β* = *β* (*t*) that varies both in space and time [13, 22]. For easier interpretation, we express the contact rate,

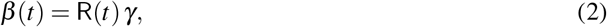

in terms of the dynamic effective reproduction number R(*t*). Here, we adopt a stochastic process approach to define R(*t*) and construct a Gaussian process latent variable model. We assume a one-mean Gaussian process prior [23],

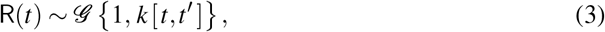

which draws function values from a multivariate normal distribution. This normal distribution is parameterized in terms of the covariance function *k* [*t,t ′*] and assumes that R(t) is constant within a time window of five days. To account for a smooth non-linear mapping from the latent to the data space, we choose an exponentiated quadratic form of the covariance function with two kernel hyperparameters, *η*^2^ and *𝓁*^2^ [6, 10].

### 2.2 Mobility modeling

We represent the disease dynamics in each state by its own local SEIR model and connect all 50 states to Stanford campus using a discrete mobility network [12]. Figure 1 shows that this network consists of *n*_nd_ = 50 + 1 nodes, which represent the individual states, and *n*_eg_ = 50 weighted edges, which represent the strength of their connection to Stanford campus. We approximate the weights of the edges by the number incoming students from each state and represent this information in the adjacency matrix *A*_*ij*_ and in the degree matrix *D*_*II*_,

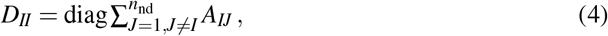

The difference between the degree matrix *D*_*IJ*_ and the adjacency matrix *A*_*IJ*_ defines the weighted graph Laplacian *L*_*IJ*_, [3, 12, 21],

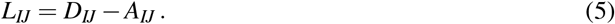

**Fig. 1.**
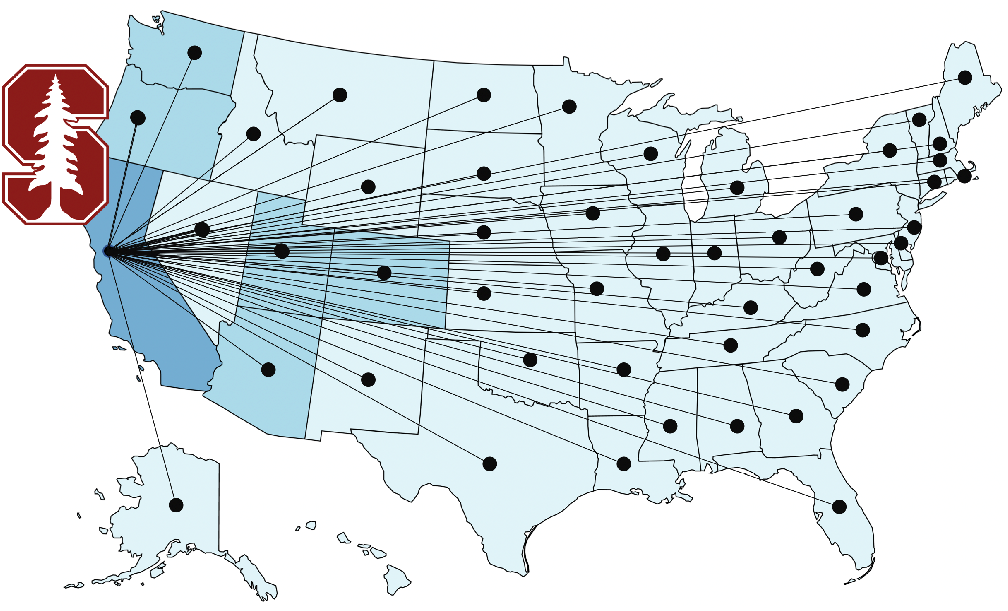
Mobility modeling. Discrete graphs of the returning student network model with *n*_nd_ = 51 nodes and *n*_eg_ = *n*_nd_ −1 edges that represent the mobility of students returning to Stanford University campus.

Since we focus on the return to campus, we only simulate the mobility to Stanford campus and neglect the intrastate mobility between individual states. This implies that only a single row and column of the adjacency matrix and degree matrix are populated [14], *A*_*I*1_ = *A*_1J_ ≠ 0 and *D*_11_ ≠ 0, while all other entries are zero, *A*_*IJ*_ = 0 for all *I, J* ≠ 1 and *D*_*II*_ = 0 for all *I* ≠1. We assume that all students arrive at the beginning of a given term rather than continuously over time. In this sense, we only use the network structure to calculate the influx of students at the first day of the term as an initial condition for the model.

### 2.3 Bayesian inference

Our model local SEIR features four parameters: two parameters that define the initial exposed and infectious populations *E*_0_ and *I*_0_, and two kernel-hyperparameters for the Gaussian process prior *η*^2^ and *l*^2^ to model the dynamics of the effective reproduction number R(*t*). This implies that our network SEIR model introduces four model parameters for each state,

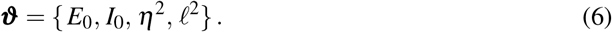

We use Bayes’ theorem to estimate the posterior probability distribution of the parameters ***ϑ***, such that the statistics of the simulated daily new cases *D*(***ϑ***, *t*) agree with the reported daily new cases 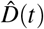,

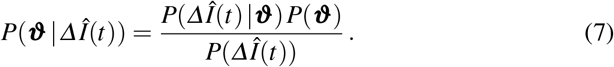

Here *P*(*ΔÎ*(*t*) ***ϑ***) is the likelihood, the conditional probability of the data *Î*(*t*) for given fixed parameters ***ϑ*** ; *P*(***ϑ***) is the prior, the probability distribution of the model parameters ***ϑ*** ; *P*(*ΔÎ*(*t*)) is the marginal likelihood; and *P*(***ϑ*** *ΔÎ*(*t*)) is the posterior, the conditional probability of parameters ***ϑ*** for given data *ΔÎ*(*t*).

#### Likelihood

The likelihood function evaluates the goodness of fit between the simulated new case numbers *ΔI*(***ϑ***, *t*) = *I*(***ϑ***, *t* + 1) − *I*(***ϑ***, *t*) and the reported new case numbers *ΔÎ*(*t*). For the individual likelihood functions *ℒ* (*ΔÎ*(*t*_*i*_) |***ϑ***), at each time point *t*_*i*_, we adopt a Student’s *t*-distribution with a case-number-dependent width,

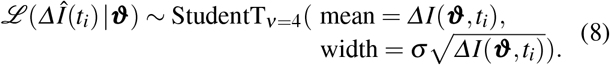

We assume a half Cauchy distribution for the likelihood width *σ* between the simulated and reported new cases *ΔI*(***ϑ***, *t*) and *ΔÎ*(*t*),

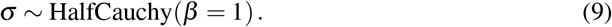

The product of all *i* = 0.., n likelihood functions *ℒ* (*ΔÎ*(*t*_*i*_) |***ϑ***), evaluated at the daily time points *t*_*i*_, defines the overall likelihood *P*(*Î*(*t*)|***ϑ***),

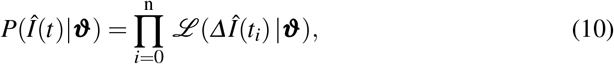

where the product symbol 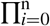 denotes the multiplication of all *i* = 0.., n daily likelihoods across the time window of the simulation.

#### Priors

For the prior probability distributions *P*(***ϑ***), we select log-normal distributions for the initial exposed and infectious populations,

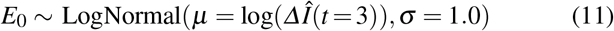

and

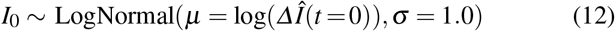

and half Cauchy and Gamma distributions for the two kernel hyperparameters that define the exponentiated quadratic form of the covariance function for the Gaussian process model,

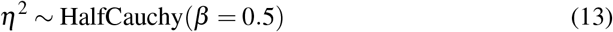

and

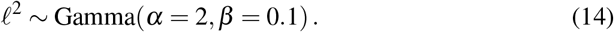

#### Posteriors

We evaluate Bayes’ theorem (7) using Markov Chain Monte Carlo sampling [13, 20, 22] to obtain the posterior distribution *P*(***ϑ*** |*ΔÎ*(*t*)) of the parameters ***ϑ*** = {*E*_0_, *I*_0_, *η*^2^, *𝓁*^2^} in terms of the likelihood *P*(*Î*(*t*) |***ϑ***) and prior probability distributions *P*(***ϑ***). We adopt the NO-U-Turn sampler [7] implemented in the Python package PyMC3 [24]. From the converged posterior distributions, we sample multiple combinations of parameters ***ϑ***. From these posterior samples, we quantify and plot the means and 95% credible intervals of the reproduction number R(*t*) and the simulated daily new cases *ΔI*(***ϑ***, *t*) for all 50 states.

### 2.4 COVID-19 outbreak and mobility data

For the COVID-19 outbreak data, we draw the COVID-19 history for all 50 states and Santa Clara County [26]. From these data, we extract the daily newly cases *Î*(*t*). To eliminate weekday-weekend fluctuations, we smoothen the outbreak data by applying a moving an averaging window of seven days. For the mobility data, we draw the student demographics of all Stanford University undergraduates enrolled in 2020, broken down by their origin state and their year of study, frosh, sophomore, junior, or senior year.

### 2.5 Campus opening forecast

We analyze the COVID-19 dynamics for campus opening at four different quarters and for the three currently most common virus variants. To model the effect of campus opening, we allow students to travel back to Stanford campus using our network SEIR model. We investigate campus opening for the spring quarter on April 6, 2020, the summer quarter on June 22, 2020, the fall quarter September 14, 2020 and the winter quarter January 11, 2021. For each quarter, we estimate the effects of opening by assuming the dynamic reproduction number R(*t*) of Santa Clara County, that we infer directly from the local COVID-19 outbreak data from Santa Clara County. To model the effects of three different virus variants, in addition to the baseline simulation, we consider the B.1.1.7 variant with an increased transmissibility of 56% and the B.1.351 variant with an increased transmissibility of 50% [4]. For both virus variants, we scale our inferred effective reproduction number of Santa Clara by multiplication with the increased transmissibility and perform forward simulations for all four quarters, spring, summer, fall, and winter.

## 3 Results

### 3.1 COVID-19 outbreak dynamics in the United States

Figure 2 illustrates the outbreak dynamics of COVID-19 in the United States from the beginning of the outbreak until January 17, 2021. For each state, the bottom graph represents the new confirmed cases *ΔI*(*t*) as dots and our inferred dynamic SEIR model fit *ΔÎ*(*t*) as orange curves. The shaded regions represent our inferred 95% credible interval on the daily new cases. The top graph represents the inferred evolution of the effective reproduction number R(*t*). The solid lines describe the median values of R(*t*) and the shaded areas represent the 95% credible intervals. The side-by-side comparison of all 50 states showcases the different disease dynamics and the varying timing of the COVID-19 waves in each state. While most states saw high effective reproduction numbers and a strong increase in new COVID-19 cases during the months of November and December 2020, the early outbreak dynamics displayed larger regional differences in waves between the individual states. These differences are of particular interest for informing the intrinsic risk that inviting students back to campus entails for a university campus like Stanford. For example, New York had a large outbreak at the beginning of the pandemic but managed to keep its effective reproduction numbers low until the end of the summer. In contrast, states like Arizona, California, Florida, Nevada, South Carolina and Texas saw a high regional COVID-19 prevalence by the end of June 2020, around the time Stanford’s summer quarter was about to start.

**Fig. 2.**
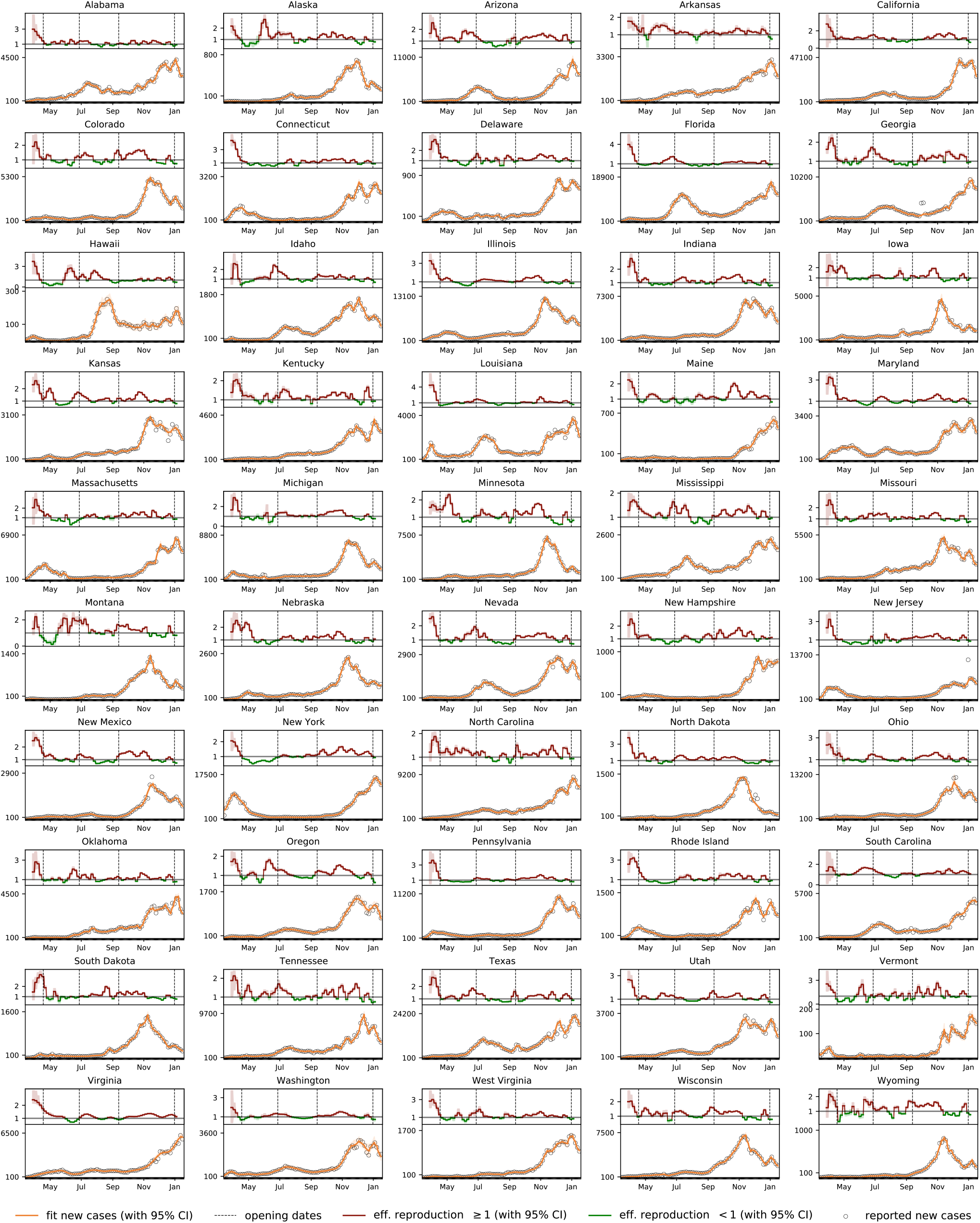
Outbreak dynamics of COVID-19 in the United States. Effective reproduction number (red and green curves) and daily new cases (circles) with model fit (orange curves) for all 50 states from the beginning of the outbreak until January 17, 2021. Solid lines represent the median values, shaded areas highlight the 95% credible intervals.

To invite undergraduates from all of the United States back to campus, we need to consider the epidemiological status of each states at the time a new quarter starts, which is showcased in Figure 3. The left plot summarizes the effective reproduction number variation and the right plot showcases the exposed and infectious population sizes throughout all the states at the beginning of each quarter. Figure 3 shows that, at the beginning of the spring, summer, and fall quarters, most states across the United States saw imminent outbreaks with effective reproduction numbers well above one. At the same time, the relative sizes of the exposed and infectious populations were still relatively small. At the start of the winter quarter, the opposite situation occurred as most states were recovering from a new COVID-19 wave. Reproduction were well below one in most states, but the relative exposed *E* and infectious *I* population sizes were still significantly larger than at the beginning of the other quarters.

**Fig. 3.**
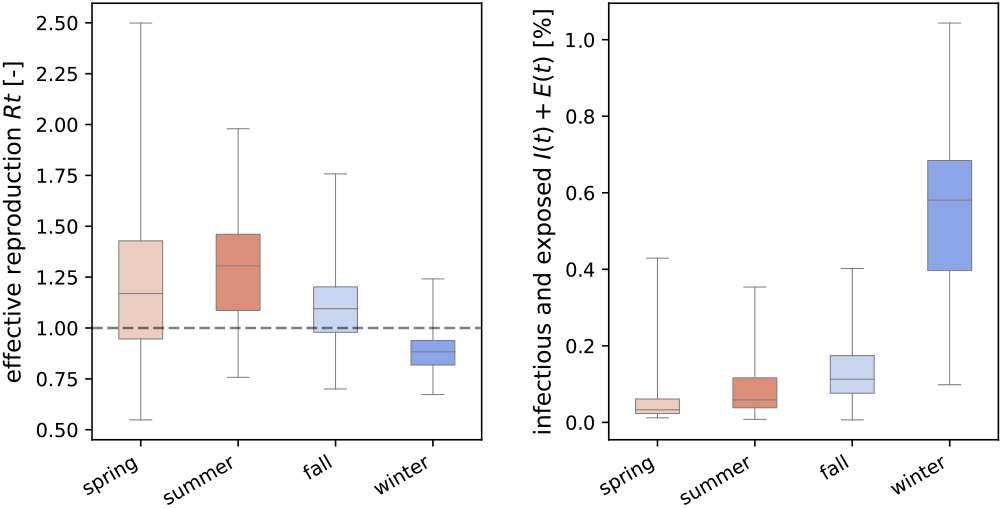
Epidemiological status of the incoming student population for four consecutive quarters. Effective reproduction statistics and size of the exposed and infectious populations at the beginning of the spring, summer, fall, and winter quarters.

For this study, these population sizes are of most interest, as they determine how many infectious students will return to campus. The spring, summer, fall, and winter quarter started on April 6, June 22, September 14, 2020 and Jan 11, 2021. Table 1 summarizes the infectious and exposed populations in each state for each of these potential campus reopening dates. On April 6, the largest exposed and infectious populations were 0.43% in New York, 0.34% in New Jersey, and 0.20% in Massachusetts. Consequently, for the spring quarter, students returning from the East Coast had the highest chances of bringing COVID-19 back to campus. On June 22, we inferred a 0.35%, 0.22% and 0.20% exposed and infectious population in Arizona, Florida, and South Carolina. On September 14, students returning from North Dakota, South Dakota, and Wisconsin had a 0.41%, 0.29% and 0.26% chance to be exposed or infectious. On Jan 11, peak exposed and infectious populations were recorded at 1.04%, 0.93%, and 0.84% in Arizona, California, and South Carolina. These numbers clearly show the significantly higher risk in inviting students back to campus at the beginning of the winter quarter: The average exposed and infectious populations across all states were 0.54% at the start of the winter quarter, compared to 0.13%, 0.08% and 0.06% for the fall, summer, and spring.

**Table 1.**
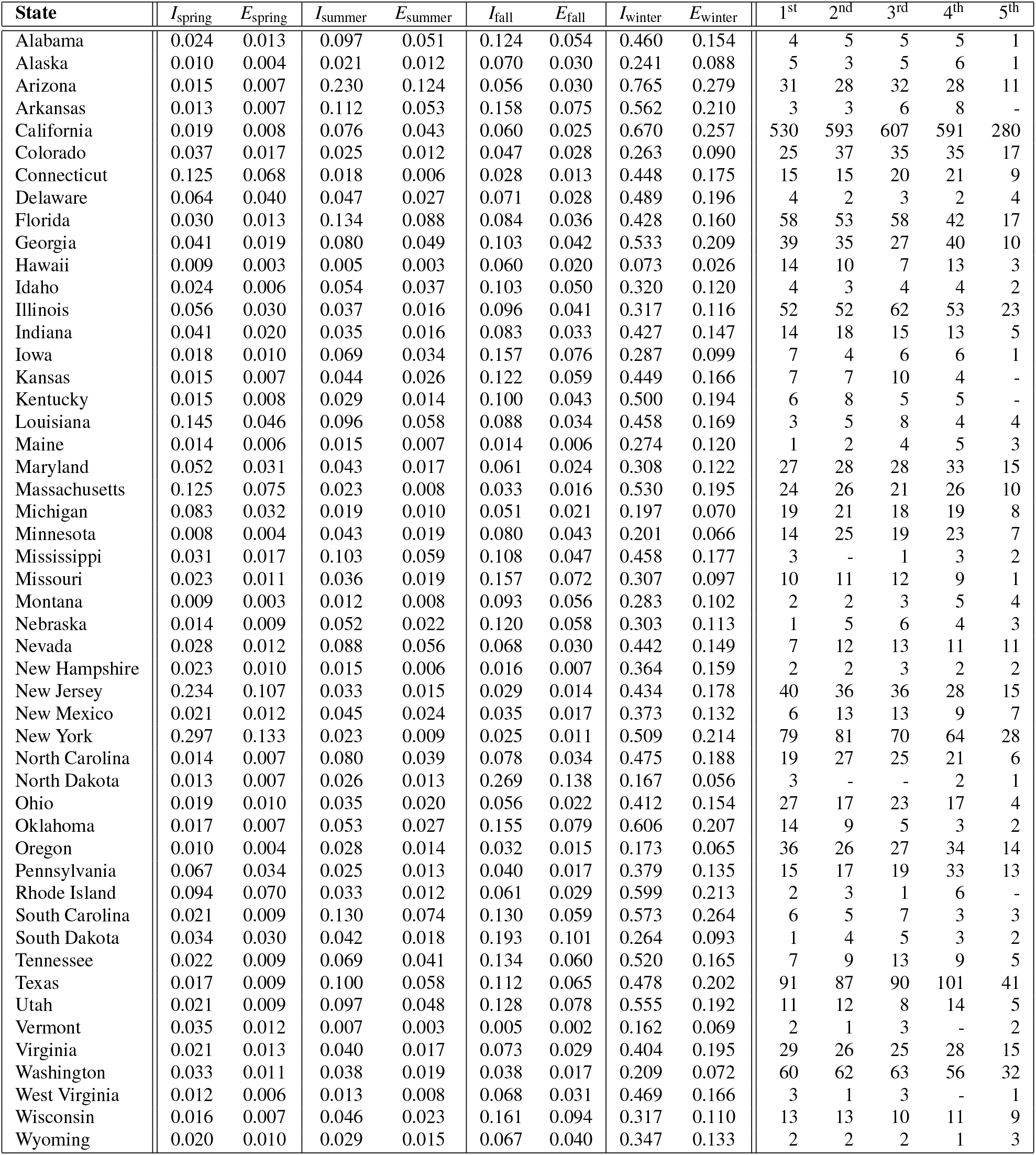
Outbreak dynamics of COVID-19 in the United States. Percentages of infectious, and exposed populations for four consecutive quarters and number of 1st, 2nd, 3rd, 4th, and 5th year undergraduates originating from each state.

| State | $I_{spring}$ | $E_{spring}$ | $I_{summer}$ | $E_{summer}$ | $I_{fall}$ | $E_{fall}$ | $I_{winter}$ | $E_{winter}$ | 1<sup>st</sup> | 2<sup>nd</sup> | 3<sup>rd</sup> | 4<sup>th</sup> | 5<sup>th</sup> |
| --- | --- | --- | --- | --- | --- | --- | --- | --- | --- | --- | --- | --- | --- |
| Alabama | 0.024 | 0.013 | 0.097 | 0.051 | 0.124 | 0.054 | 0.460 | 0.154 | 4 | 5 | 5 | 5 | 1 |
| Alaska | 0.010 | 0.004 | 0.021 | 0.012 | 0.070 | 0.030 | 0.241 | 0.088 | 5 | 3 | 5 | 6 | 1 |
| Arizona | 0.015 | 0.007 | 0.230 | 0.124 | 0.056 | 0.030 | 0.765 | 0.279 | 31 | 28 | 32 | 28 | 11 |
| Arkansas | 0.013 | 0.007 | 0.112 | 0.053 | 0.158 | 0.075 | 0.562 | 0.210 | 3 | 3 | 6 | 8 | - |
| California | 0.019 | 0.008 | 0.076 | 0.043 | 0.060 | 0.025 | 0.670 | 0.257 | 530 | 593 | 607 | 591 | 280 |
| Colorado | 0.037 | 0.017 | 0.025 | 0.012 | 0.047 | 0.028 | 0.263 | 0.090 | 25 | 37 | 35 | 35 | 17 |
| Connecticut | 0.125 | 0.068 | 0.018 | 0.006 | 0.028 | 0.013 | 0.448 | 0.175 | 15 | 15 | 20 | 21 | 9 |
| Delaware | 0.064 | 0.040 | 0.047 | 0.027 | 0.071 | 0.028 | 0.489 | 0.196 | 4 | 2 | 3 | 2 | 4 |
| Florida | 0.030 | 0.013 | 0.134 | 0.088 | 0.084 | 0.036 | 0.428 | 0.160 | 58 | 53 | 58 | 42 | 17 |
| Georgia | 0.041 | 0.019 | 0.080 | 0.049 | 0.103 | 0.042 | 0.533 | 0.209 | 39 | 35 | 27 | 40 | 10 |
| Hawaii | 0.009 | 0.003 | 0.005 | 0.003 | 0.060 | 0.020 | 0.073 | 0.026 | 14 | 10 | 7 | 13 | 3 |
| Idaho | 0.024 | 0.006 | 0.054 | 0.037 | 0.103 | 0.050 | 0.320 | 0.120 | 4 | 3 | 4 | 4 | 2 |
| Illinois | 0.056 | 0.030 | 0.037 | 0.016 | 0.096 | 0.041 | 0.317 | 0.116 | 52 | 52 | 62 | 53 | 23 |
| Indiana | 0.041 | 0.020 | 0.035 | 0.016 | 0.083 | 0.033 | 0.427 | 0.147 | 14 | 18 | 15 | 13 | 5 |
| Iowa | 0.018 | 0.010 | 0.069 | 0.034 | 0.157 | 0.076 | 0.287 | 0.099 | 7 | 4 | 6 | 6 | 1 |
| Kansas | 0.015 | 0.007 | 0.044 | 0.026 | 0.122 | 0.059 | 0.449 | 0.166 | 7 | 7 | 10 | 4 | - |
| Kentucky | 0.015 | 0.008 | 0.029 | 0.014 | 0.100 | 0.043 | 0.500 | 0.194 | 6 | 8 | 5 | 5 | - |
| Louisiana | 0.145 | 0.046 | 0.096 | 0.058 | 0.088 | 0.034 | 0.458 | 0.169 | 3 | 5 | 8 | 4 | 4 |
| Maine | 0.014 | 0.006 | 0.015 | 0.007 | 0.014 | 0.006 | 0.274 | 0.120 | 1 | 2 | 4 | 5 | 3 |
| Maryland | 0.052 | 0.031 | 0.043 | 0.017 | 0.061 | 0.024 | 0.308 | 0.122 | 27 | 28 | 28 | 33 | 15 |
| Massachusetts | 0.125 | 0.075 | 0.023 | 0.008 | 0.033 | 0.016 | 0.530 | 0.195 | 24 | 26 | 21 | 26 | 10 |
| Michigan | 0.083 | 0.032 | 0.019 | 0.010 | 0.051 | 0.021 | 0.197 | 0.070 | 19 | 21 | 18 | 19 | 8 |
| Minnesota | 0.008 | 0.004 | 0.043 | 0.019 | 0.080 | 0.043 | 0.201 | 0.066 | 14 | 25 | 19 | 23 | 7 |
| Mississippi | 0.031 | 0.017 | 0.103 | 0.059 | 0.108 | 0.047 | 0.458 | 0.177 | 3 | - | 1 | 3 | 2 |
| Missouri | 0.023 | 0.011 | 0.036 | 0.019 | 0.157 | 0.072 | 0.307 | 0.097 | 10 | 11 | 12 | 9 | 1 |
| Montana | 0.009 | 0.003 | 0.012 | 0.008 | 0.093 | 0.056 | 0.283 | 0.102 | 2 | 2 | 3 | 5 | 4 |
| Nebraska | 0.014 | 0.009 | 0.052 | 0.022 | 0.120 | 0.058 | 0.303 | 0.113 | 1 | 5 | 6 | 4 | 3 |
| Nevada | 0.028 | 0.012 | 0.088 | 0.056 | 0.068 | 0.030 | 0.442 | 0.149 | 7 | 12 | 13 | 11 | 11 |
| New Hampshire | 0.023 | 0.010 | 0.015 | 0.006 | 0.016 | 0.007 | 0.364 | 0.159 | 2 | 2 | 3 | 2 | 2 |
| New Jersey | 0.234 | 0.107 | 0.033 | 0.015 | 0.029 | 0.014 | 0.434 | 0.178 | 40 | 36 | 36 | 28 | 15 |
| New Mexico | 0.021 | 0.012 | 0.045 | 0.024 | 0.035 | 0.017 | 0.373 | 0.132 | 6 | 13 | 13 | 9 | 7 |
| New York | 0.297 | 0.133 | 0.023 | 0.009 | 0.025 | 0.011 | 0.509 | 0.214 | 79 | 81 | 70 | 64 | 28 |
| North Carolina | 0.014 | 0.007 | 0.080 | 0.039 | 0.078 | 0.034 | 0.475 | 0.188 | 19 | 27 | 25 | 21 | 6 |
| North Dakota | 0.013 | 0.007 | 0.026 | 0.013 | 0.269 | 0.138 | 0.167 | 0.056 | 3 | - | - | 2 | 1 |
| Ohio | 0.019 | 0.010 | 0.035 | 0.020 | 0.056 | 0.022 | 0.412 | 0.154 | 27 | 17 | 23 | 17 | 4 |
| Oklahoma | 0.017 | 0.007 | 0.053 | 0.027 | 0.155 | 0.079 | 0.606 | 0.207 | 14 | 9 | 5 | 3 | 2 |
| Oregon | 0.010 | 0.004 | 0.028 | 0.014 | 0.032 | 0.015 | 0.173 | 0.065 | 36 | 26 | 27 | 34 | 14 |
| Pennsylvania | 0.067 | 0.034 | 0.025 | 0.013 | 0.040 | 0.017 | 0.379 | 0.135 | 15 | 17 | 19 | 33 | 13 |
| Rhode Island | 0.094 | 0.070 | 0.033 | 0.012 | 0.061 | 0.029 | 0.599 | 0.213 | 2 | 3 | 1 | 6 | - |
| South Carolina | 0.021 | 0.009 | 0.130 | 0.074 | 0.130 | 0.059 | 0.573 | 0.264 | 6 | 5 | 7 | 3 | 3 |
| South Dakota | 0.034 | 0.030 | 0.042 | 0.018 | 0.193 | 0.101 | 0.264 | 0.093 | 1 | 4 | 5 | 3 | 2 |
| Tennessee | 0.022 | 0.009 | 0.069 | 0.041 | 0.134 | 0.060 | 0.520 | 0.165 | 7 | 9 | 13 | 9 | 5 |
| Texas | 0.017 | 0.009 | 0.100 | 0.058 | 0.112 | 0.065 | 0.478 | 0.202 | 91 | 87 | 90 | 101 | 41 |
| Utah | 0.021 | 0.009 | 0.097 | 0.048 | 0.128 | 0.078 | 0.555 | 0.192 | 11 | 12 | 8 | 14 | 5 |
| Vermont | 0.035 | 0.012 | 0.007 | 0.003 | 0.005 | 0.002 | 0.162 | 0.069 | 2 | 1 | 3 | - | 2 |
| Virginia | 0.021 | 0.013 | 0.040 | 0.017 | 0.073 | 0.029 | 0.404 | 0.195 | 29 | 26 | 25 | 28 | 15 |
| Washington | 0.033 | 0.011 | 0.038 | 0.019 | 0.038 | 0.017 | 0.209 | 0.072 | 60 | 62 | 63 | 56 | 32 |
| West Virginia | 0.012 | 0.006 | 0.013 | 0.008 | 0.068 | 0.031 | 0.469 | 0.166 | 3 | 1 | 3 | - | 1 |
| Wisconsin | 0.016 | 0.007 | 0.046 | 0.023 | 0.161 | 0.094 | 0.317 | 0.110 | 13 | 13 | 10 | 11 | 9 |
| Wyoming | 0.020 | 0.010 | 0.029 | 0.015 | 0.067 | 0.040 | 0.347 | 0.133 | 2 | 2 | 2 | 1 | 3 |

### 3.2 COVID-19 influx into Stanford campus

The risk of bringing students back to campus depends on both the regional covid prevalence and the state-specific origin of the returning student population. The last five columns of Table 1 report the origin of Stanford University’s current undergraduate population. Figure 4 showcases the inferred amount of exposed and infectious students returning to campus from each state. We estimate the COVID-19 influx by multiplying the state-specific prevalence with the number of students returning from each state, and summarize these values in Table 2. For a potential opening in the spring quarter, we inferred one COVID-19 exposed or infectious student returning from New York, one from California, and one from New Jersey. For a summer opening, our results suggest that we could have expected three exposed or infectious students returning from California and one exposed or infectious student from Texas and Florida. For a fall opening, we would have seen two exposed or infectious individuals returning from California and one from Texas. At the beginning of the winter quarter, the high COVID-19 prevalence throughout the United States would have resulted in 24 exposed or infectious students returning from within the state, three from Texas, two from New York, and one from Arizona, Florida, Georgia, Illinois, New Jersey, Massachusetts, Washington, Virginia, North Carolina, Maryland, and Colorado each.

**Fig. 4.**
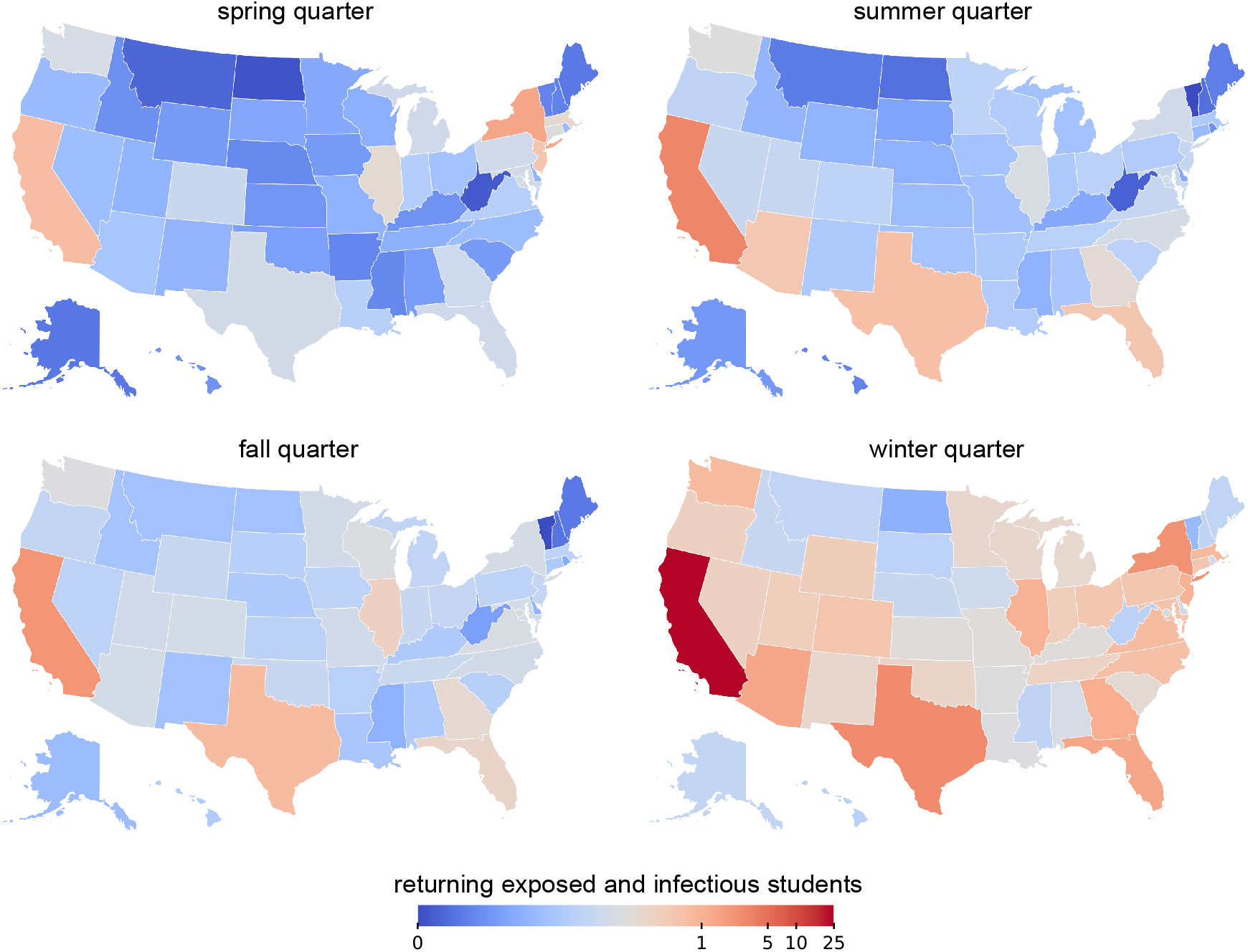
COVID-19 influx into Stanford campus. Incoming infectious and exposed students from the United States at the beginning of the spring, summer, fall, and winter quarters.

**Table 2.** COVID-19 influx into Stanford campus. Incoming infectious and exposed students from the United States at the beginning of the spring, summer, fall, and winter quarters.

| States | spring | summer | fall | winter | enrolled |
| --- | --- | --- | --- | --- | --- |
| Alabama | 0 | 0 | 0 | 0 | 20 |
| Alaska | 0 | 0 | 0 | 0 | 20 |
| Arizona | 0 | 0 | 0 | 1 | 130 |
| Arkansas | 0 | 0 | 0 | 0 | 20 |
| California | 1 | 3 | 2 | 24 | 2601 |
| Colorado | 0 | 0 | 0 | 1 | 149 |
| Connecticut | 0 | 0 | 0 | 0 | 80 |
| Delaware | 0 | 0 | 0 | 0 | 15 |
| Florida | 0 | 1 | 0 | 1 | 228 |
| Georgia | 0 | 0 | 0 | 1 | 151 |
| Hawaii | 0 | 0 | 0 | 0 | 47 |
| Idaho | 0 | 0 | 0 | 0 | 17 |
| Illinois | 0 | 0 | 0 | 1 | 242 |
| Indiana | 0 | 0 | 0 | 0 | 65 |
| Iowa | 0 | 0 | 0 | 0 | 24 |
| Kansas | 0 | 0 | 0 | 0 | 28 |
| Kentucky | 0 | 0 | 0 | 0 | 24 |
| Louisiana | 0 | 0 | 0 | 0 | 24 |
| Maine | 0 | 0 | 0 | 0 | 15 |
| Maryland | 0 | 0 | 0 | 1 | 131 |
| Massachusetts | 0 | 0 | 0 | 1 | 107 |
| Michigan | 0 | 0 | 0 | 0 | 85 |
| Minnesota | 0 | 0 | 0 | 0 | 88 |
| Mississippi | 0 | 0 | 0 | 0 | 9 |
| Missouri | 0 | 0 | 0 | 0 | 43 |
| Montana | 0 | 0 | 0 | 0 | 16 |
| Nebraska | 0 | 0 | 0 | 0 | 19 |
| Nevada | 0 | 0 | 0 | 0 | 54 |
| New Hampshire | 0 | 0 | 0 | 0 | 11 |
| New Jersey | 1 | 0 | 0 | 1 | 155 |
| New Mexico | 0 | 0 | 0 | 0 | 48 |
| New York | 1 | 0 | 0 | 2 | 322 |
| North Carolina | 0 | 0 | 0 | 1 | 98 |
| North Dakota | 0 | 0 | 0 | 0 | 6 |
| Ohio | 0 | 0 | 0 | 0 | 88 |
| Oklahoma | 0 | 0 | 0 | 0 | 33 |
| Oregon | 0 | 0 | 0 | 0 | 137 |
| Pennsylvania | 0 | 0 | 0 | 0 | 97 |
| Rhode Island | 0 | 0 | 0 | 0 | 12 |
| South Carolina | 0 | 0 | 0 | 0 | 24 |
| South Dakota | 0 | 0 | 0 | 0 | 15 |
| Tennessee | 0 | 0 | 0 | 0 | 43 |
| Texas | 0 | 1 | 1 | 3 | 410 |
| Utah | 0 | 0 | 0 | 0 | 50 |
| Vermont | 0 | 0 | 0 | 0 | 8 |
| Virginia | 0 | 0 | 0 | 1 | 123 |
| Washington | 0 | 0 | 0 | 1 | 273 |
| West Virginia | 0 | 0 | 0 | 0 | 8 |
| Wisconsin | 0 | 0 | 0 | 0 | 56 |
| Wyoming | 0 | 0 | 0 | 0 | 10 |
| Total | 4 | 7 | 6 | 46 | 6479 |

### 3.3 Local outbreak dynamics in Santa Clara County

Figure 5 illustrates the local outbreak dynamics for Santa Clara County, home of Stanford University. The bottom graph depicts the reported daily new cases *ΔI*(*t*) as dots and our inferred dynamic SEIR model fit *ΔÎ*(*t*) as orange curve. The shaded regions highlight the inferred 95% credible interval on the new confirmed cases. The top graph showcases the inferred temporal evolution of the effective reproduction number R(*t*) in Santa Clara County, which serves as the input to our forward predictions in the following example. The solid lines describe the median values of R(*t*) and the shaded area depicts the 95% credible interval. Similar to the state of California in Figure 2, Santa Clara County saw two significant COVID-19 prevalence waves, the first at the beginning of July and the second from early November to the beginning of December.

**Fig. 5.**
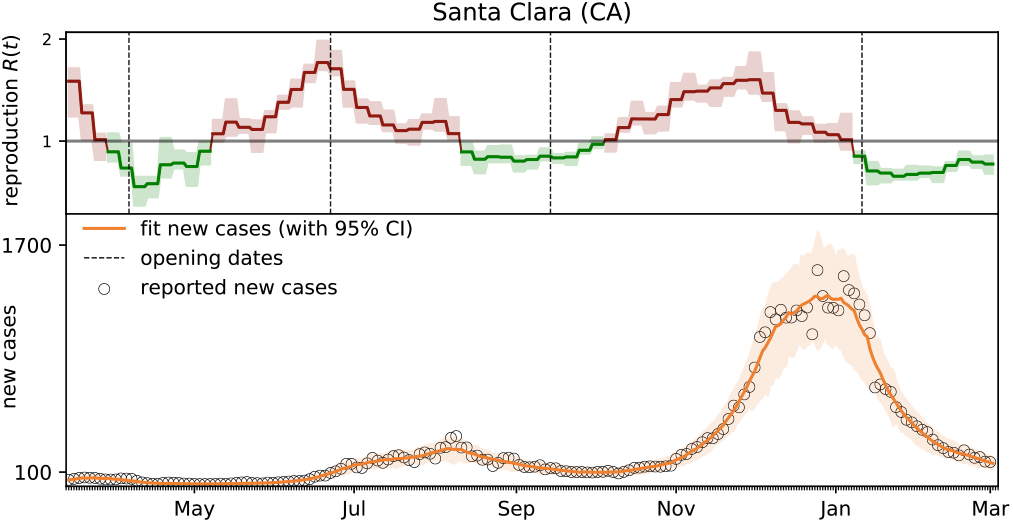
Local outbreak dynamics of COVID-19 in Santa Clara County. Effective reproduction number (red and green curves) and daily new cases (circles) with model fit (orange curves) for Santa Clara County from the beginning of the outbreak until March 3, 2021. Solid lines represent the median values, shaded areas highlight the 95% credible intervals.

### 3.4 Risk assessment of new B.1.1.7 and B.1.351 variants

Figures 6, 7 and 8 summarize our forward risk analysis for reopening campus under three different transmissibility scenarios: the true outbreak dynamics under the wild-type SARS-CoV-2, and the hypothetical outbreak dynamics under the new COVID-19 variants B.1.1.7 and B.1.351 with 56% and 50% increased transmissibility.

**Fig. 6.**
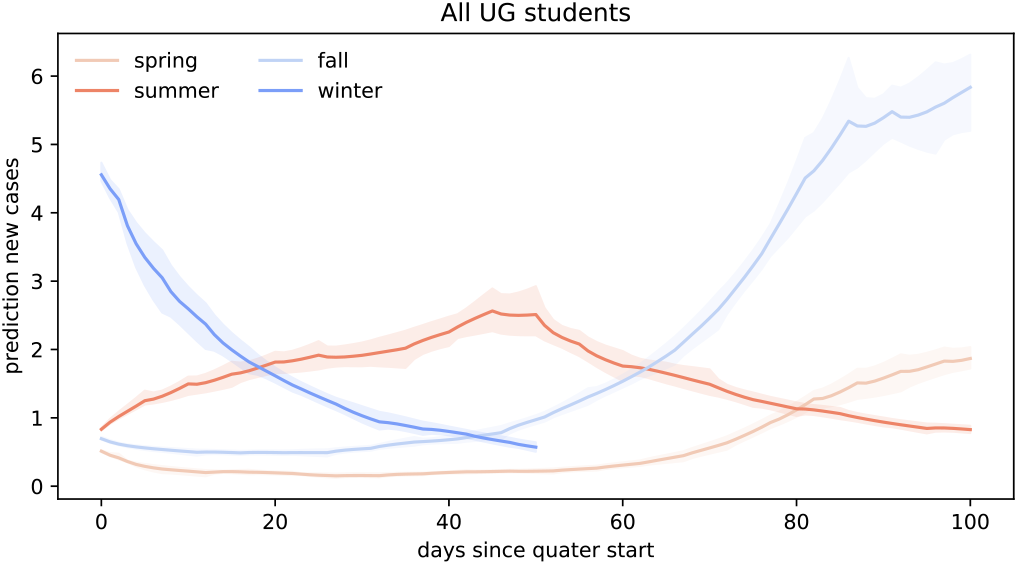
Forward prediction of new COVID cases for the wild-type SARS-CoV-2. Each simulation begins with the number of returning exposed and infectious cases on the first day of class and uses the inferred effective reproduction number R(*t*) of Santa Clara County for the forward prediction throughout the entire quarter.

**Fig. 7.**
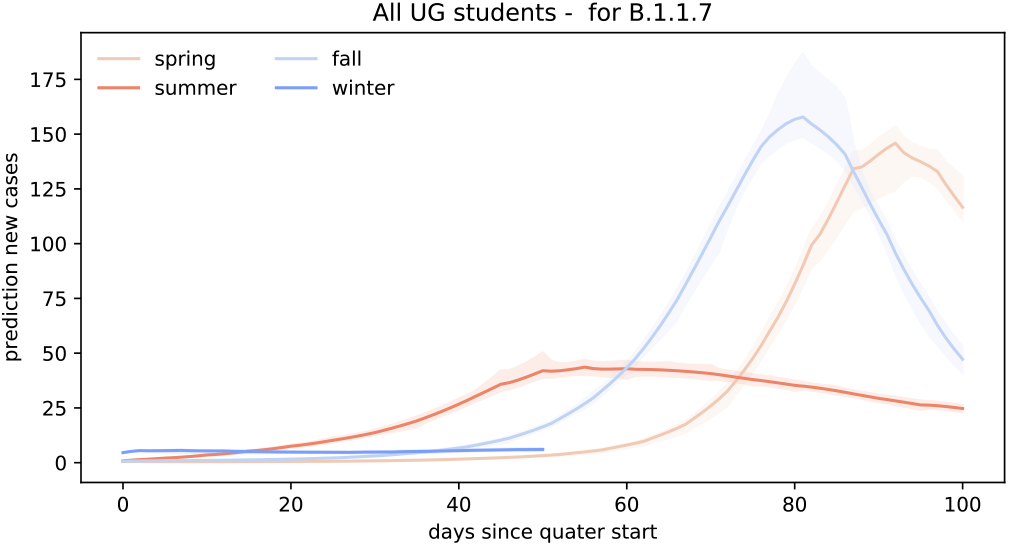
Forward prediction of new COVID cases for the B.1.1.7 variant. Each simulation begins with the number of returning exposed and infectious cases on the first day of class and uses the inferred effective reproduction number R(*t*) of Santa Clara County, scaled by the 56% higher infectiousness of B.1.1.7, for the forward prediction throughout the entire quarter.

**Fig. 8.**
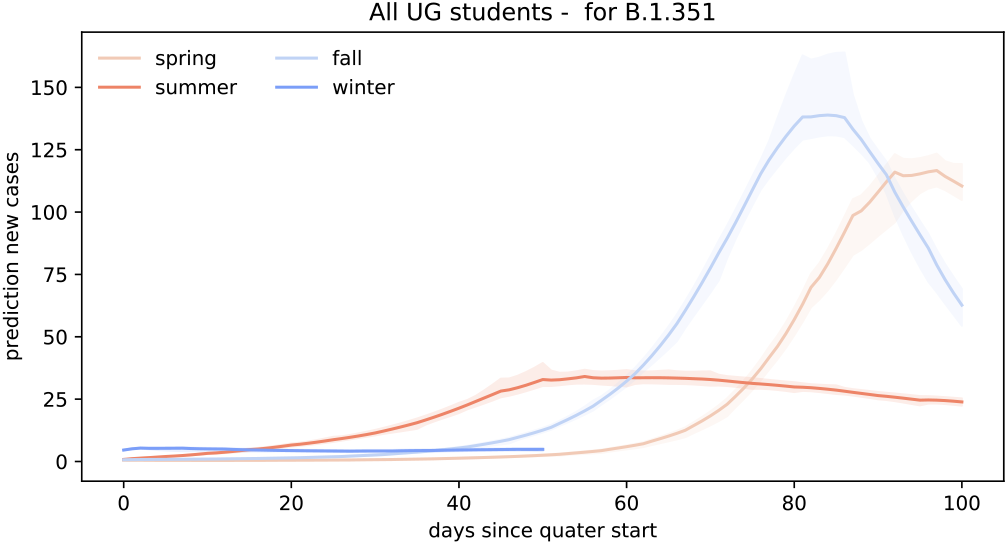
Forward prediction of new COVID cases for the B.1.351 variant. Each simulation begins with the number of returning exposed and infectious cases on the first day of class and uses the inferred effective reproduction number R(*t*) of Santa Clara County, scaled by the 50% higher infectiousness of B.1.351, for the forward prediction throughout the entire quarter.

Figure 6 represents a scenario for the wild-type SARS-CoV-2, where no new COVID-19 virus variants are brought into campus by the returning students. In this scenario, the amount of new cases per day within the first 100 days after campus reopening remains limited to a maximum of two students if Stanford University reopened in the spring of 2020. For the same scenario, we would expect a maximum of three, six, and five cases per day after reopening campus for the summer, fall, and winter quarters. In total, reopening for spring, summer, or fall quarter would have resulted in 58 cases, 162 cases, and 203 cases during the first 100 days after reopening. For reopening campus at the beginning of the winter quarter, a total of 86 cases would have to be expected by March 3, 2021.

Figure 7 represents the scenario where returning students bring in the new B.1.1.7 COVID variant which has an increased transmissibility of 56%. In this scenario, our forward predictions estimate significantly higher daily confirmed case numbers compared to the previous scenario. If students would have brought in the new B.1.1.7 COVID variant during spring quarter campus reopening, we would expect to see a significant outbreak amounting to 146 new cases per day on day 92 after reopening. Bringing students back campus during the summer quarter under the B.1.1.7 COVID-19 variant scenario would have led to a more flattened outbreak curve peaking at 44 new daily cases on day 55. Reopening Stanford University for the fall quarter with the new B.1.1.7. variant circulating on campus would have led to a steep outbreak wave reaching a maximum amount of 158 new confirmed cases on day 81. Finally, reopening in winter would have generated a slow and controlled spreading. Within the first 52 days, the maximum amount of daily new daily cases would have amounted to 6 on day 50. In total, reopening campus under the reproduction dynamics of the new B.1.1.7 variant would have led to 3329, 2555, and 4727 cases 100 days after reopening for the spring, summer, and fall quarters.

Figure 8 represents a similar scenario in which we assume that the new B.1.351. variant would be the dominant variant in the undergraduate population. Similar to the B.1.1.7 scenario, an increased transmissibility would have resulted in a steep wave when reopening campus at the beginning of the spring and fall quarters. More specifically, we could have expected 117 new cases on day 97 and 139 new cases on day 84 for the spring and fall quarters. A more flattened COVID-19 wave upon summer reopening would have peaked at 34 daily new cases on day 55. For winter quarter reopening, the amount of new cases would have stayed relatively constant at approximately four to five new cases per day during the first 52 days of the quarter. In total, the B.1.351 variant scenario predicts 2581, 2117, and 4256 cases 100 days after reopening for the spring, summer, and fall quarters and 237 cases 52 days after reopening for the winter quarter.

## 4 Conclusion

The COVID-19 pandemic remains an enormous challenge for higher education, especially for residential college campuses with a diverse student population from all across the country. Most institutions have cautiously limited student access to campus throughout the first year of the pandemic, but are now gradually inviting their students back to campus. In view of massive immunization campaigns, it seems natural to assume that it would now be safe to now transition back to in person instruction. However, since the early outbreak of the COVID-19 pandemic in December 2019, new and more contagious variants of the virus have emerged and COVID-19 is now beginning to spread predominantly into younger age groups. Taken together, these events bring back the question about safe campus opening.

In this study, we explore the effects of campus reopening for the example of Stanford University, a residential campus in Santa Clara County, California, with an undergraduate enrollment of 6479 students. For four consecutive quarters, spring, summer, fall, and winter, we virtually open Stanford campus and invite the undergraduate population from all 50 United States to return to campus under the current outbreak conditions of their home states. We use a Bayesian analysis to infer the disease parameters of each state using reported case data and extract the exposed and infectious populations at the first day of class for all four quarters. We find that 4, 7, 6, and 46 infected students would have brought the SARS-CoV-2 virus to campus if Stanford had opened in spring, summer, fall, and winter.

With these initial conditions, we perform a forward analysis and compare three scenarios: the true outbreak dynamics under the initial wild-type SARS-CoV-2, and the hypothetical outbreak dynamics under the new B.1.1.7 and B.1.351 variants with 56% and 50% increased transmissibility. We assume that the local outbreak dynamics on Stanford campus are driven by the effective reproduction number of Santa Clara County that we infer from the counties reported case data using Bayesian inference. Our study suggests that, with no additional countermeasures, the most affected quarter would have been the fall of 2020, with 203 cases under baseline reproduction, and 4727 and 4256 cases for the B.1.1.7 and B.1.351 variants.

Strikingly, the outbreak dynamics of the B.1.1.7 and B.1.351 variants are significantly different from those of the wild-type SARS-CoV-2 version. In fall, the wild-type curve increases monotonically until the end of the quarter. In contrast, for both variants, the outbreak curves peak towards the last third of the quarter, with 158 daily new cases for B.1.1.7 and 139 for B.1.351, and then steadily decline. This suggests that both variants are sufficiently infectious to create a state of herd immunity: A large enough group of students, 4727 for B.1.1.7 and 4256 for B.1.351, recover from infection throughout the fall quarter to protect the community from further spreading, although the reproduction number remains well above one. While these case numbers are certainly only an upper limit, since we neglect additional countermeasures and test-trace-isolate strategies, they clearly highlight the effects of more infectious variants of the virus.

Infectious disease experts agree that it is highly likely that more infectious variants of the virus will emerge in the near future. Our study shows that, even if a new variant is only a few percent more contagious than the wild-type version of SARS-CoV-2, this can have tremendous consequences on the overall case numbers, and, ultimately, on the health care system. College campus administrators will need to maintain tight outbreak measures, quarantine, and isolation, to continue to successfully manage their local outbreak dynamics as more campuses reopen towards the fall.

## Data Availability

Data are available on the public COVID-19 databases cited in the bibliography

## Acknowledgements

This work was supported by the Stanford School of Engineering COVID-19 Research and Assistance Fund and by a Stanford Bio-X IIP seed grant to Ellen Kuhl, by a DAAD Fellowship to Kevin Linka, by a BAEF Fellowship to Mathias Peirlinck, and by the Engineering and Physical Sciences Research Council grant EP/R020205/1 and a RAMP Continuity Network: Scientific Meetings, Rapid Review Group, and Policy Support for COVID-19 grant from the UKRI ID 108556 to Alain Goriely.

